# Dismantling cognitive-behavioral therapy for chronic insomnia: a protocol for a systematic review and component network meta-analysis

**DOI:** 10.1101/2022.06.02.22275890

**Authors:** Yuki Furukawa, Masatsugu Sakata, Satoshi Funada, Shino Kikuchi, Toshi A. Furukawa, Edoardo G. Ostinelli, Orestis Efthimiou, Michael Perlis

## Abstract

**Introduction:** Insomnia is highly prevalent and disabling. Clinical practice guidelines recommend cognitive-behavioral therapy for insomnia (CBTI) as the first-line treatment. However, CBTI includes various combinations of many different components and its clinical benefits have been shown as a package, whereas the effect of each component remains unclear. In this study, we will explore the effect of each component of CBTI with the use of component network meta-analysis.

**Methods and analysis:** We will include all randomized controlled trials that compared any form of CBTI against another form of CBTI or a control condition in the treatment of adults with chronic insomnia. Concomitant treatments will be allowed as long as they are equally distributed among the arms. We will include both primary and secondary insomnia. The primary outcome of interest in this study is (1) treatment efficacy (remission defined as reaching a satisfactory state at endpoint measured by any validated self-reported scale) at four weeks post-treatment or at its closest time point. Secondary outcomes are (2) acceptability, (3) sleep diary measures and (4) efficacy at long-term follow-up. We will systematically search in PubMed, CENTRAL, PsycINFO and WHO International Clinical Trials Registry Platform. We will assess risk of bias using Cochrane Risk of Bias 2.0 tool. We will conduct component network meta-analysis with the *netmeta* package in R.

**Ethics and dissemination:** This study will use published data and does not require ethical approval. Findings will be disseminated in a peer-reviewed journal.

**PROSPERO:** CRD42022324233.

## REVIEW QUESTION

What is the effect of each component of cognitive behavioral therapy for chronic insomnia?

## BACKGROUND

Insomnia is highly prevalent and disabling.(Roth et al., 2011) Clinical practice guidelines recommend cognitive-behavioral therapy for insomnia (CBTI) as the first-line treatment.(Edinger et al., 2021; Qaseem et al., 2016) CBTI includes various combinations of many different components and its clinical benefits have been shown as a package,(Straten et al., 2018) whereas the effect of each component remains unclear. Finding the effect of each component will lead to an intervention that may maximize treatment benefit while reducing the number of components, thereby reducing the treatment burden, lowering the cost of CBTI and making it available to more people. Component network meta-analysis (CNMA) is an extension of standard network meta-analysis that can be used to disentangle the treatment effects of different components included in multicomponent interventions.(Rücker et al., 2020b) In this study, we will explore the effect of each component of CBTI with the use of CNMA.

## METHODS

We will follow the Preferred Reporting Items for Systematic reviews and Meta-Analyses (PRISMA) guideline extension for NMA. (Hutton et al., 2015) The protocol is prospectively registered in PROSPERO (CRD42022324233).

### Data sources

#### Criteria for considering studies for this review

##### Study design

We will include all randomized controlled trials that compared any form of CBTI against another form of CBTI or a control condition in the treatment of adults with chronic insomnia. Cluster randomized trials will be included in meta-analyses in accordance with the Cochrane handbook recommendation.(Higgins et al., 2022) If the intra-cluster correlation coefficient is not clearly reported, we assume it to be 0.05.(Lin et al., 2018)

##### Participants

We will include studies on patients of both genders aged 18 years or older with insomnia either diagnosed according to formal diagnostic criteria (such as the Diagnostic and Statistical Manual of Mental Disorders, the International Classification of Diseases or the International Classification of Sleep Disorders) or judged so by clinical expertise (e.g. presence of significant symptoms). The effect of including studies without a formal diagnosis of insomnia will be tested in a sensitivity analysis. We will include patients with psychiatric or physical comorbidities.(Sateia, 2014) The effect of including such studies will be examined in a sensitivity analysis.

##### Interventions and controls

We regard CBTI broadly as a psychotherapy involving any one of the following cognitive or behavioral components. Table 1 describes the different components of interest and their definitions. Pharmacological or psychological co-administration will be allowed as long as it is stated that there were no systematic differences between study arms. CBTI that clearly includes active components focusing on other symptoms, such as depression, anxiety or pain will be excluded when such components are not equally distributed among study arms.

**TABLE 1.** List of included components and their definitions.

| **Intervention** | **Description** |
| --- | --- |
| **Educational components (EDU)** |  |
| Sleep hygiene education (se) | General explanation about sleep (eg, sleep biology, characteristics of healthy sleep, changes in sleep patterns with aging, stress biology, and its impact on sleep) and general recommendations about lifestyle (eg, diet, exercise, substance use) and environmental factors (eg, light, noise, temperature) to improve sleep. This may include some elements of other components but those should not be the predominant part of the intervention. |
| Sleep diary (sd) | Self-monitoring of important daily sleep-related information using diary. |
| **Cognitive components (COG)** |  |
| Cognitive restructuring (cr) | Skills to identify, challenge and change unhelpful beliefs about sleep that may disturb sleep. This may include behavioral experiments. |
| Mindfulness (mi) | A form of meditation emphasizing a nonjudgmental state of heightened or complete awareness of one’s thoughts, emotions, or experiences on a moment-to-moment basis. |
| Constructive worry (cw) | Skills not to worry in bed by writing down the worries and their solutions before going to bed. |
| Imagery rehearsal (ir) | Skills to identify, challenge and change nightmares that disturb sleep. |
| **Behavioral components (BEH)** |  |
| Sleep restriction (sr) | Skills to improve sleep by limiting time in bed. First, time in bed is restricted to the average sleep duration and then it is increased or decreased depending on sleep efficiency. |
| Stimulus control (sc) | Skills to re-associate the bed with sleep. Patients are instructed to; wake up at the same time every morning, refrain from daytime napping, go to bed only when sleepy, get out of bed when unable to sleep, and use the bed/bedroom for sleep and sex only. |
| Relaxation (re) | Structured exercises designed to reduce somatic tension (eg, abdominal breathing, progressive muscle relaxation, autogenic training) and cognitive arousal (eg, guided imagery training). |
| Paradoxical intention (pi) | Exercise to remain awake as long as possible after getting into bed. |
| **Others** |  |
| Waiting component (w) | Participants are aware that they can receive an active treatment after a waiting phase. If patients allocated to the waiting list control condition receive some other potentially therapeutic components, we will consider both the waiting component and the therapeutic components to be present. |
| Conventional drug treatment (dt) | Rated positive when conventional drug treatment is present (drug treatment is part of the protocol treatment) or allowed (we will note the percentage of patients on drug). |
| Non-specific treatment effect (ns) | Effect of an intervention due to the patients' belief that they are receiving some form of treatment. Miscellaneous skills not covered in other sections and not expected to have large effect (i.e. quasi-desensitization) are classified as having non-specific treatment effect. |
| Self-help, unguided, remote (NA) | Default delivery format is self-help, unguided and remote, as it requires the least human resource. |
| Human encouragement (he) | Reminders provided by human beings to proceed with the self-help, remote treatment program via telephone or email. This should not contain any support related to the therapeutic contents. Peer support such as discussion group will be regarded as this component. We will code this component separately from interaction with therapists (in, gp, ff) to see if adding human encouragement to self-help, remote interventions good enough to be effective. |
| Therapeutic guidance (tg) | Therapeutic guidance in addition to self-help, remote interventions. This may be provided on a scheduled basis or as-needed basis. Technical support only is not included. We will code this component separately from interaction with therapists (in, gp, ff) to see if adding therapeutic guidance to self-help, remote interventions good enough to be effective. |
| Individual (in) | Individual interaction with therapists. The combination (in + ff) means individual, face-to-face sessions are held. The combination (in – ff) individual remote sessions, such as via telephone or videoconference. |
| Group (gp) | Interaction with therapists as a member of a group. |
| Face-to-face (ff) | Face-to-face interaction with therapists. |
| Automatic encouragement (ae) | Automated reminders to proceed with the treatment program. This can be added both to the self-help interventions or the interventions including interactions with therapists. This should not contain any support related to the therapeutic contents. |

The control conditions of interest will include waiting list, no treatment, attention/psychological placebo control and treatment as usual. In this CNMA study, treatment as usual must include pharmacotherapy; watchful waiting will be classified as attention/psychological placebo even when it is termed ‘treatment as usual’ in some papers. When no treatment, attention/psychological placebo or treatment as usual are used while on the waiting list, such controls will be regarded as waiting list in the NMA but will be decomposed as appropriate in the CNMA. We will include interventions of any duration.

Where multiple arms are reported in a single trial, we will include only the relevant arms that can be described with the components listed in Table 1. We will lump the arms when they share the same components (e.g. same components delivered in 30 min per session against 60 min per session). We will exclude irrelevant arms such as active-drug, pill-placebo, exercise or bright light therapy with equipment. Possible components and their combinations are shown in Table 1 and Table 2. The components and nodes were determined based on previous studies (Edinger et al., 2021; Furukawa et al., 2021) and content expert consensus among co-authors (MS, TAF, MP).

**TABLE 2.**
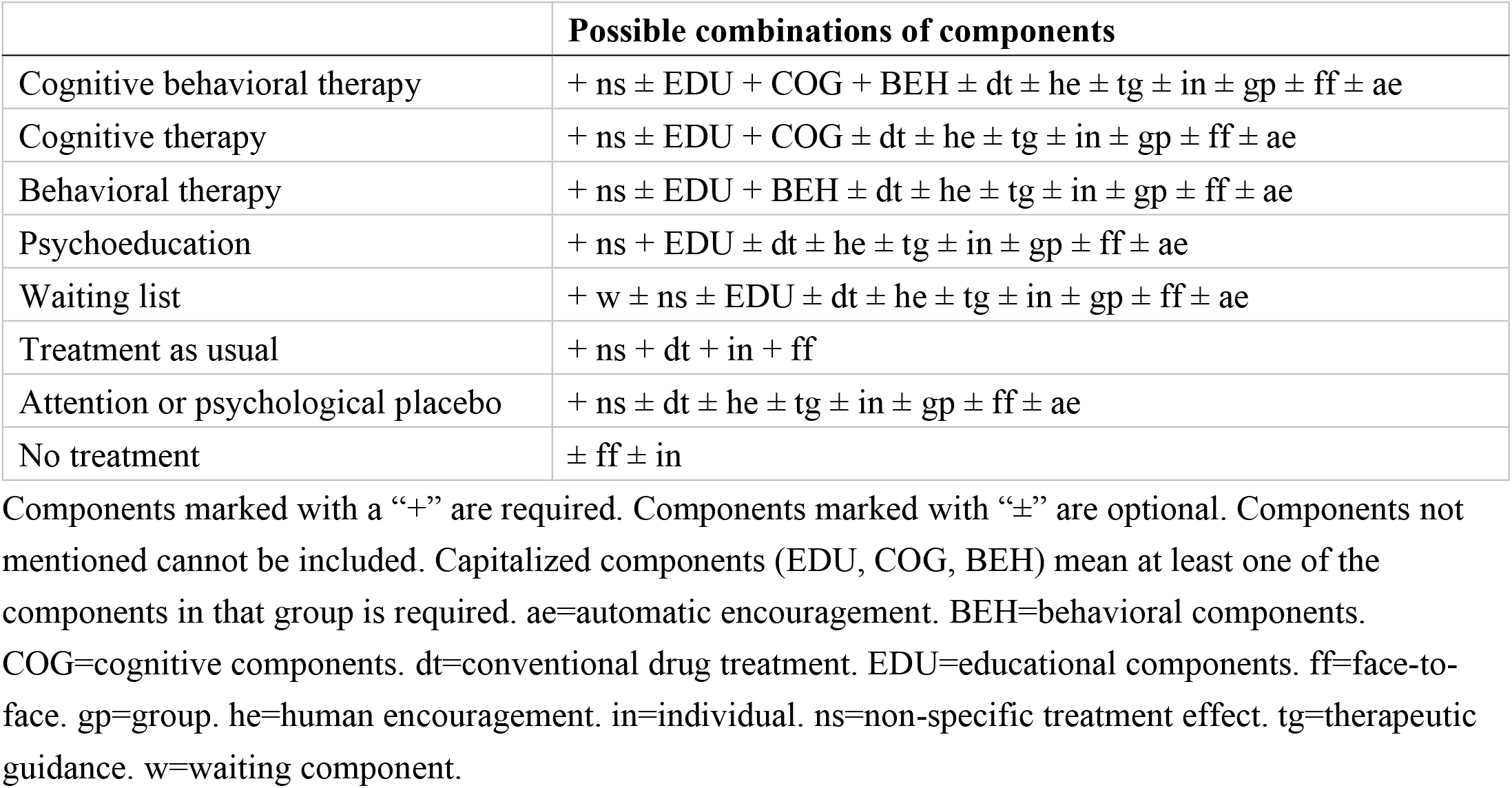
Conceptualization of cognitive behavioral therapy for insomnia or control conditions from the component perspective.

|  | **Possible combinations of components** |
| --- | --- |
| Cognitive behavioral therapy | + ns ± EDU + COG + BEH ± dt ± he ± tg ± in ± gp ± ff ± ae |
| Cognitive therapy | + ns ± EDU + COG ± dt ± he ± tg ± in ± gp ± ff ± ae |
| Behavioral therapy | + ns ± EDU + BEH ± dt ± he ± tg ± in ± gp ± ff ± ae |
| Psychoeducation | + ns + EDU ± dt ± he ± tg ± in ± gp ± ff ± ae |
| Waiting list | + w ± ns ± EDU ± dt ± he ± tg ± in ± gp ± ff ± ae |
| Treatment as usual | + ns + dt + in + ff |
| Attention or psychological placebo | + ns ± dt ± he ± tg ± in ± gp ± ff ± ae |
| No treatment | ± ff ± in |

Components marked with a “+” are required. Components marked with “±” are optional. Components not mentioned cannot be included. Capitalized components (EDU, COG, BEH) mean at least one of the components in that group is required. ae=automatic encouragement. BEH=behavioral components. COG=cognitive components. dt=conventional drug treatment. EDU=educational components. ff=face-to-face. gp=group. he=human encouragement. in=individual. ns=non-specific treatment effect. tg=therapeutic guidance. w=waiting component.

**TABLE 3.**
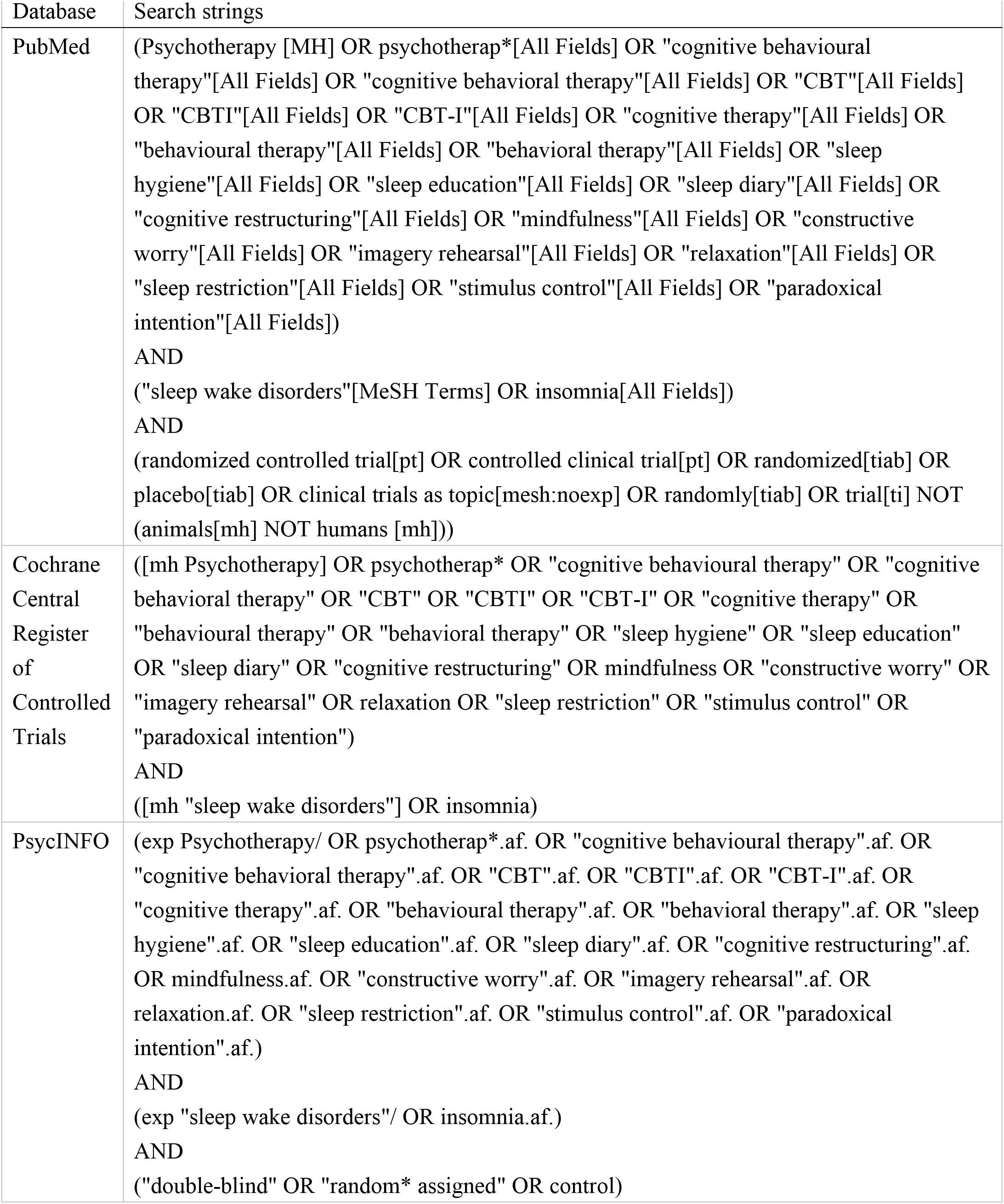
Search strings for PubMed, Cochrane Central Register of Controlled Trials and PsycINFO.

### Search methods for identification of studies

We will carry out a comprehensive literature search in PubMed, CENTRAL and PsycINFO. We will use a combination of index and free terms of psychological treatments and insomnia with filters (Eady et al., 2008) for randomized clinical trials. We will also search WHO International Clinical Trials Registry Platform. We will impose no date, language or publication status restriction. We will check the reference lists of review articles for additional potentially eligible records.

### Data collection and analysis

#### Selection of studies

Two review authors will independently screen titles and abstracts of all the potential studies we identify as a result of the search and code them as ‘retrieve’ or ‘do not retrieve’. We will retrieve the full text study reports/publications and two review authors will independently screen the full text and identify studies for inclusion and identify and record reasons for exclusion of the ineligible studies. We will resolve any disagreement through discussion or, if required, we will consult a third reviewer. We will identify publications from the same study so that each study rather than each report is the unit of analysis in the review. We will record the selection process in sufficient detail to complete a PRISMA flow diagram.

#### Data items

Two review authors will extract independently data from the included studies. Any disagreement will be resolved through discussion, or discussed with a third person if necessary. We will abstract the following information.

##### 1. Characteristics of the studies

Name of the study, year of publication, country, study site (single or multi-center), study design (individually-randomized or cluster-randomized), population characteristics (mean age, number of women, definition of insomnia, number of patients with primary insomnia), intervention (components, delivery format, qualification of therapists [if applicable], duration), outcomes (scale used for the primary outcome)

##### 2. Identification of components

Two independent reviewers will determine the classification of all identified arms and their components according to the definitions in Table 1, based on all available information including the publications, trial registry and inquiry with the original investigators if needed. Any disagreement will be solved by the two reviewers and, where necessary, in consultation with a third member of the review team. We will report the inter-rater agreement in terms of percentage agreement and kappa.

##### 3. Risk of bias

We will use Cochrane Risk of Bias 2.0 tool (RoB2) (Sterne et al., 2019) to assess the risk of bias of the primary outcome. We will report the inter-rater agreement in terms of percentage agreement and kappa.

##### 4. Data to calculate effect sizes

We will extract data to calculate effect sizes (the number of patients randomized to each arm, the number of patients assessed, the number of remitters, the scale used, the mean, standard deviation and the number assessed for continuous outcomes) When only change from baseline to endpoint is reported for continuous outcomes, we will use it instead of endpoint mean.(Costa et al., 2013)

### Primary outcome and secondary outcomes

The primary outcome of interest in this study is treatment efficacy at four weeks post-treatment or at its closest time point. Intention-to-treat analysis will be prioritized whenever available. We will use the number of participants randomized as the denominator for dichotomous outcomes. We will use odds ratio for dichotomous outcomes, mean difference for continuous outcomes expressed in minutes and percent.

1. Efficacy: remission defined as reaching a satisfactory state at endpoint measured by any validated self-reported scale (dichotomous)

Secondary outcomes are as follows;

2. Acceptability: dropouts for any reason (dichotomous)

3. Sleep diary measures (continuous)

3.1. Sleep efficiency (SE, %)

3.2. Total sleep time (TST, min)

3.3. Sleep onset latency (SOL, min)

3.4. Wake after sleep onset (WASO, min)

4. Efficacy at long-term follow-up. (dichotomous, longest follow-up between 3 to 12 months)

### Hierarchy of outcome measures

For efficacy, we will prioritize the remission using the Insomnia Severity Index (7 or less points at endpoint) (Morin et al., 2011) and its imputed number. If it is not reported, we will use the following scales in this order: the remission using the Functional Outcomes of Sleep Questionnaire-10 (18 or more points at endpoint) (Weaver et al., 2007), and then its imputed number; remission using the Epworth Sleepiness Scale (10 or less points at endpoint) (Johns, 1990), and then its imputed number; remission using the Pittsburgh Sleep Quality Index (5 or less points at baseline) (Cole et al., 2006), and then its imputed number; remission using the Athens Insomnia Scale (5 or less points at endpoint) (Okajima et al., 2020), and then its imputed number; remission using any other validated self-reported scales; remission using sleep diary measures (both SOL and WASO less than 30 minutes at endpoint. When SOL and WASO are reported only separately, we will prioritize WASO. When only SOL is reported, we will use SOL.) (Edinger et al., 2009; American Academy of Sleep Medicine, 2014) and its imputed number.

When any of the measure is reported using another definition of remission than stated above, we will use the definition stated by the authors. When any of the measure is reported only in continuous values, we will impute remission using mean and standard deviation. We will test the validity of this imputation method using studies that report the outcome both in continuous and dichotomous manner. When any of the measure is reported only in standardized mean difference, we will convert it into odds ratio using a validated method.(Chinn, 2000)

### Statistical analysis

We will perform the analysis in *R* (latest version, R foundation, Vienna, Austria) (R Core Team, 2020) using *netmeta* (latest version) package (Rücker et al., 2020a) to conduct component network meta-analysis and *meta* (latest version) package (Balduzzi et al., 2019) to synthesize the outcomes in placebo arms and to assess the publication bias.

We will first perform a network meta-analysis lumping arms that include both cognitive and behavioral components as “cognitive-behavioral therapy,” those that involve cognitive but not behavioral components as “cognitive therapy” and those that involve behavioral but not cognitive components as “behavioral therapy” to gain a first insight of the relative treatment effects. (Table 2) We will examine the transitivity assumption by creating a table of important trial and patient characteristics to see if potential effect modifiers (publication year, proportion of patients with primary insomnia, age) are similarly distributed among comparisons. We will check the consistency of the network using local and global inconsistency tests.

Then we will perform component-level network meta-analysis. We will use a model that assumes additivity of components, i.e. assuming that the effect of combination therapy is the sum of the effects of its components. Given the expected clinical and methodological heterogeneity of treatment effects among the studies, we will use the random-effects model.

### Certainty of evidence

We will assess the certainty of evidence in network estimates of the primary outcome using CINeMA.(Nikolakopoulou et al., 2020)

### Publication bias

We will assess the presence of small study effects, including publication bias, in the evidence set by examining asymmetry in the contour-enhanced funnel plots of all active interventions (CBT, CT, BT) vs control condition (psychoeducation, waiting list, treatment as usual, attention or psychological placebo, no treatment) using the primary outcome.

### Sensitivity analyses

We will conduct sensitivity analyses using the primary outcome.

1. Excluding studies without formal diagnosis of insomnia
2. Excluding studies focusing on patients with comorbidities (both physical and psychological)
3. Excluding studies with overall high dropout rate (20% or more)
4. Excluding studies at high overall risk of bias
5. Using completer-set analysis (dichotomous)

### Patient and public involvement

There was no patient or public involvement in the development of this manuscript.

## Data Availability

Data and code used for analyses will be available from the corresponding author on reasonable request.

## Acknowledgements

The views expressed are those of the authors and not necessarily those of affiliated organizations.

## Registration

This protocol is prospectively registered in PROSPERO (CRD42022324233).

This research was prospectively registered (#2022033NIe), Ethical Committee, Faculty of Medicine, The University of Tokyo.

## Changes from the protocol (since the first registration on PROSPERO)

25^th^ May, 2022 (during screening, before data extraction). Although we stated in the protocol that “CBTI that clearly includes active components focusing on other symptoms, such as depression, anxiety or pain, will be excluded,” in case the same intervention component was included in both arms, we decided to include the trial, as we will be able to see the additional benefit of CBTI.

## Contributions of authors

Conceiving the protocol: YF, MS, TAF

Designing the protocol: YF, MS, SF, SK, TAF, EGO, OE, MP

Coordinating the protocol: YF

Designing search strategies: YF, EGO

Writing the protocol: YF

Providing general advice on the protocol: TAF, OE, MP

Providing advice on statistical analysis: OE

## Support

No financial support was used.

## Declarations of interest

YF has received consultancy fee from Panasonic outside the submitted work.

MS reports personal fees from SONY outside the submitted work.

SF has a research grant from JSPS KAKENHI Grant Number JP 20K18964 and the KDDI Foundation. SK has a research grant from Mental Health Okamoto Memorial Foundation and Fujiwara Memorial Foundation.

TAF reports grants and personal fees from Mitsubishi-Tanabe, personal fees from SONY, grants and personal fees from Shionogi, outside the submitted work; In addition, TAF has a patent 2020-548587 concerning smartphone CBT apps pending, and intellectual properties for Kokoro-app licensed to Mitsubishi-Tanabe.

EGO has received research and consultancy fees from Angelini Pharma. EGO is supported by the National Institute for Health Research (NIHR) Research Professorship to Professor Andrea Cipriani (grant RP-2017-08-ST2-006), by the National Institute for Health Research (NIHR) Applied Research Collaboration (ARC) Oxford and Thames Valley, by the National Institute for Health Research (NIHR) Oxford cognitive health Clinical Research Facility and by the NIHR Oxford Health Biomedical Research Centre (grant BRC-1215-20005)

OE was supported by the Swiss National Science Foundation (Ambizione grant number 180083) MP reports no competing interest.

## Notes

### Clinical Protocols

https://www.crd.york.ac.uk/prospero/display_record.php?RecordID=324233

### Author Declarations

The study will use ONLY publicly available data located by our systematic review.

## REFERENCE

American Academy of Sleep Medicine. 2014. International Classification of Sleep Disorders.

Balduzzi S, Rücker G, Schwarzer G. How to perform a meta-analysis with R: a practical tutorial. Évid Based Ment Heal 2019;22:153. https://doi.org/10.1136/ebmental-2019-300117.

Chinn S. A simple method for converting an odds ratio to effect size for use in meta-analysis. Stat Med 2000;19:3127–31. https://doi.org/10.1002/1097-0258(20001130)19:22<3127::aid-sim784>3.0.co;2-m.

Cole JC, Motivala SJ, Buysse DJ, Oxman MN, Levin MJ, Irwin MR. Validation of a 3-Factor Scoring Model for the Pittsburgh Sleep Quality Index in Older Adults. Sleep 2006;29:112–6. https://doi.org/10.1093/sleep/29.1.112.

Costa BR da, Nüesch E, Rutjes AW, Johnston BC, Reichenbach S, Trelle S, et al. Combining follow-up and change data is valid in meta-analyses of continuous outcomes: a meta-epidemiological study. J Clin Epidemiol 2013;66:847–55. https://doi.org/10.1016/j.jclinepi.2013.03.009.

Eady AM, Wilczynski NL, Haynes RB, Team for the H. PsycINFO search strategies identified methodologically sound therapy studies and review articles for use by clinicians and researchers. J Clin Epidemiol 2008;61:34–40. https://doi.org/10.1016/j.jclinepi.2006.09.016.

Edinger JD, Arnedt JT, Bertisch SM, Carney CE, Harrington JJ, Lichstein KL, et al. Behavioral and psychological treatments for chronic insomnia disorder in adults: an American Academy of Sleep Medicine clinical practice guideline. J Clin Sleep Med 2021;17:255–62. https://doi.org/10.5664/jcsm.8986.

Edinger JD, Olsen MK, Stechuchak KM, Means MK, Lineberger MD, Kirby A, et al. Cognitive Behavioral Therapy for Patients with Primary Insomnia or Insomnia Associated Predominantly with Mixed Psychiatric Disorders: a Randomized Clinical Trial. Sleep 2009;32:499–510. https://doi.org/10.1093/sleep/32.4.499.

Furukawa TA, Suganuma A, Ostinelli EG, Andersson G, Beevers CG, Shumake J, et al. Dismantling, optimising, and personalising internet cognitive behavioural therapy for depression: a systematic review and component network meta-analysis using individual participant data. Lancet Psychiatry 2021;8:500–11. https://doi.org/10.1016/s2215-0366(21)00077-8.

Higgins J, Eldridge S, Li T. Chapter 23: Including variants on randomized trials. In: Higgins J, Eldridge S, Li T, editors. Cochrane Handbook for Systematic Reviews of Interventions version 6.3, 2022.

Hutton B, Salanti G, Caldwell DM, Chaimani A, Schmid CH, Cameron C, et al. The PRISMA Extension Statement for Reporting of Systematic Reviews Incorporating Network Meta-analyses of Health Care Interventions: Checklist and Explanations. Ann Intern Med 2015;162:777–84. https://doi.org/10.7326/m14-2385.

Johns MW. A New Method for Measuring Daytime Sleepiness: The Epworth Sleepiness Scale. Sleep 1990;14:540–5. https://doi.org/10.1093/sleep/14.6.540.

Lin C-Y, Strong C, Scott AJ, Broström A, Pakpour AH, Webb TL. A cluster randomized controlled trial of a theory-based sleep hygiene intervention for adolescents. Sleep 2018;41. https://doi.org/10.1093/sleep/zsy170.

Morin CM, Belleville G, Bélanger L, Ivers H. The Insomnia Severity Index: Psychometric Indicators to Detect Insomnia Cases and Evaluate Treatment Response. Sleep 2011;34:601–8. https://doi.org/10.1093/sleep/34.5.601.

Nikolakopoulou A, Higgins JPT, Papakonstantinou T, Chaimani A, Giovane CD, Egger M, et al. CINeMA: An approach for assessing confidence in the results of a network meta-analysis. Plos Med 2020;17:e1003082. https://doi.org/10.1371/journal.pmed.1003082.

Okajima I, Miyamoto T, Ubara A, Omichi C, Matsuda A, Sumi Y, et al. Evaluation of Severity Levels of the Athens Insomnia Scale Based on the Criterion of Insomnia Severity Index. Int J Environ Res Pu 2020;17:8789. https://doi.org/10.3390/ijerph17238789.

Qaseem A, Kansagara D, Forciea MA, Cooke M, Denberg TD, Physicians CGC of the AC of. Management of Chronic Insomnia Disorder in Adults: A Clinical Practice Guideline From the American College of Physicians. Ann Intern Med 2016;165:125. https://doi.org/10.7326/m15-2175.

R Core Team. R: A language and environment for statistical computing. R Foundation for Statistical Computing. 2020.

Roth T, Coulouvrat C, Hajak G, Lakoma MD, Sampson NA, Shahly V, et al. Prevalence and Perceived Health Associated with Insomnia Based on DSM-IV-TR; International Statistical Classification of Diseases and Related Health Problems, Tenth Revision; and Research Diagnostic Criteria/International Classification of Sleep Disorders, Second Edition Criteria: Results from the America Insomnia Survey. Biol Psychiat 2011;69:592–600. https://doi.org/10.1016/j.biopsych.2010.10.023.

Rücker G, Krahn U, König J, Efthimiou O, Schwarzer G. netmeta: Network Meta-Analysis using Frequentist Methods. 2020a.

Rücker G, Petropoulou M, Schwarzer G. Network meta-analysis of multicomponent interventions. Biometrical J Biometrische Zeitschrift 2020b;62:808–21. https://doi.org/10.1002/bimj.201800167.

Sateia MJ. International Classification of Sleep Disorders-Third Edition. Chest 2014;146:1387–94. https://doi.org/10.1378/chest.14-0970.

Sterne JA, Savović J, Page MJ, Elbers RG, Blencowe NS, Boutron I, et al. RoB 2: a revised tool for assessing risk of bias in randomised trials. Bmj 2019;366:l4898. https://doi.org/10.1136/bmj.l4898.

Straten A van, Zweerde T van der, Kleiboer A, Cuijpers P, Morin CM, Lancee J. Cognitive and behavioral therapies in the treatment of insomnia: A meta-analysis. Sleep Med Rev 2018;38:3–16. https://doi.org/10.1016/j.smrv.2017.02.001.

Weaver TE, Maislin G, Dinges DF, Bloxham T, George CFP, Greenberg H, et al. Relationship Between Hours of CPAP Use and Achieving Normal Levels of Sleepiness and Daily Functioning. Sleep 2007;30:711–9. https://doi.org/10.1093/sleep/30.6.711.

